# A practical technique for clinicians to approximate platelet function after fifteen-minutes using thromboelastography

**DOI:** 10.1101/2020.03.10.20033860

**Authors:** Elliott Sharp, Vanessa Fludder

## Abstract

**Background:** Point-of-care platelet function tests are used by anaesthetists and surgeons to create risk management plans for patients who have recently taken antiplatelet medication. Thromboelastography (TEG), one method of determining platelet function, sometimes takes >60 minutes to produce results. Previous studies have shown a novel parameter, area under the curve at 15 minutes (AUC15), correlates with clinical outcomes but used privately-owned, custom-made software to calculate AUC15. This study aimed to create a formula that clinicians can use to approximate AUC15 which correlates to the most widely used measure of platelet function, percentage of platelet aggregation.

**Methods:** Platelet function after 15 minutes can be approximated by the equation: *AUC15 =* 225(tan(θ)) where θ = α(MA_ADP_/MA_Thrombin_). A retrospective database review was performed on eligible platelet function tests that assessed ADP receptor inhibition on a TEG 6s Hemostasis Analyzer (Haemonetics^®^) across 15 months. Results were analysed using a bivariate scatter plot with linear regression line and a two-tailed Pearson correlation coefficient was calculated.

**Results:** Forty-seven tests were retrieved, of which, forty-five were eligible for analysis. Pearson two-tailed correlation coefficient showed that AUC15 correlated significantly with percentage of platelet aggregation (R = 0.748, 95% CI [0.582, 0.854], p < 0.001).

**Conclusion:** This study creates the first practical method for clinicians to approximate platelet function on TEG analysers after 15 minutes, instead of >60 minutes, using routinely generated outputs and a calculator. Clinicians who use this method will afford themselves more time to create risk management plans for patients which may improve patient outcomes.

## 1. INTRODUCTION

Point-of-care platelet function tests (PFTs) are clinician performed, non-laboratory tests that stimulate platelet aggregation, assess the time taken until a clot is formed and the strength of that clot. Current UK guidelines recommend that patients stop taking all adenosine diphosphate (ADP) receptor antagonists (clopidogrel, prasugrel and ticagrelor) a minimum of 5 days prior to an operation and 7 days prior to neuraxial anaesthesia.[1] However, in emergencies, patients may have taken an ADP receptor antagonist within the last 24 hours and be at an increased risk of suffering haemorrhagic complications.

PFTs are frequently used to assess the intra-operative bleeding risk of a patient taking antiplatelet drugs in need of an emergency surgical procedure.[2]

PFTs can also guide the anaesthetic plan of multimorbid patients taking antiplatelets. In these patients, neuraxial anaesthesia may be preferred over general anaesthesia because of the risk of increased morbidity from a general approach. However, patients who are taking ADP receptor antagonists may be at an increased risk of neuraxial haematoma formation from neuraxial anaesthesia and subsequent neurological damage.[3] PFTs can help determine the degree of platelet inhibition in these patients and may allow anaesthetists to make an informed choice about their anaesthetic plan when used in conjunction with evidence about clinical outcomes in neuraxial anaesthesia, such as the National Audit Project on Major Complications of Central Neuraxial Blocks (NAP3).[4]

Time is often a scarce resource in both of these emergency scenarios where a patient has not stopped their ADP receptor antagonist for the recommended time prior to needing an emergency operation or neuraxial anaesthesia. Thromboelastography (TEG), is a relatively widely used point-of-care PFT that measures viscoelastic changes to blood to determine the inhibition of platelet P2Y_12_ receptors of ADP.[5] However, some TEG results can take upwards of 60 minutes to be reported which is a problem if more urgent surgical or anaesthetic decisions are required.[6]

A previous study has validated a novel parameter, the area under the thromboelastograph curve after 15 minutes (AUC15), to rapidly assess an individual’s *ex vivo* clotting response to clopidogrel, an ADP receptor antagonist, in healthy volunteers and patients undergoing percutaneous coronary intervention (PCI).[6,7] However, the authors calculated AUC15 using a, “specific [computer] program designed and written for [their] study”.[7] As of the writing of this article, the custom-made computer programme used in this study has not been released for public use. Even it was to be publicly released, existing TEG analysers would have to be modified to install the new software. Therefore, a practical technique that all clinicians can use is required to calculate the AUC15. Since existing TEG analysers do not typically provide exact equations of the curves, the available limited parameters must be used to approximate AUC15 instead. To the knowledge of authors, no published method exists for clinicians to calculate the AUC15 for the TEG ADP curve.

### 1.1. Aims

The aim of this study is to create a formula that clinicians can use to reliably approximate the area AUC15 for TEG platelet function tests to determine the *ex vivo* inhibition of platelet ADP receptors.

## 2. EQUIPMENT AND METHODS

All anonymised PFTs which assessed ADP receptor function conducted at a large teaching hospital on a TEG 6s Hemostasis Analyzer (Haemonetics^®^) across 15 months were retrieved. For the TEG 6s analyser, these tests are also known as the Platelet Mapping™ assay. An example of a typical TEG 6s Platelet Mapping™ tracing can be seen in Figure 1a.

**Figure 1.**
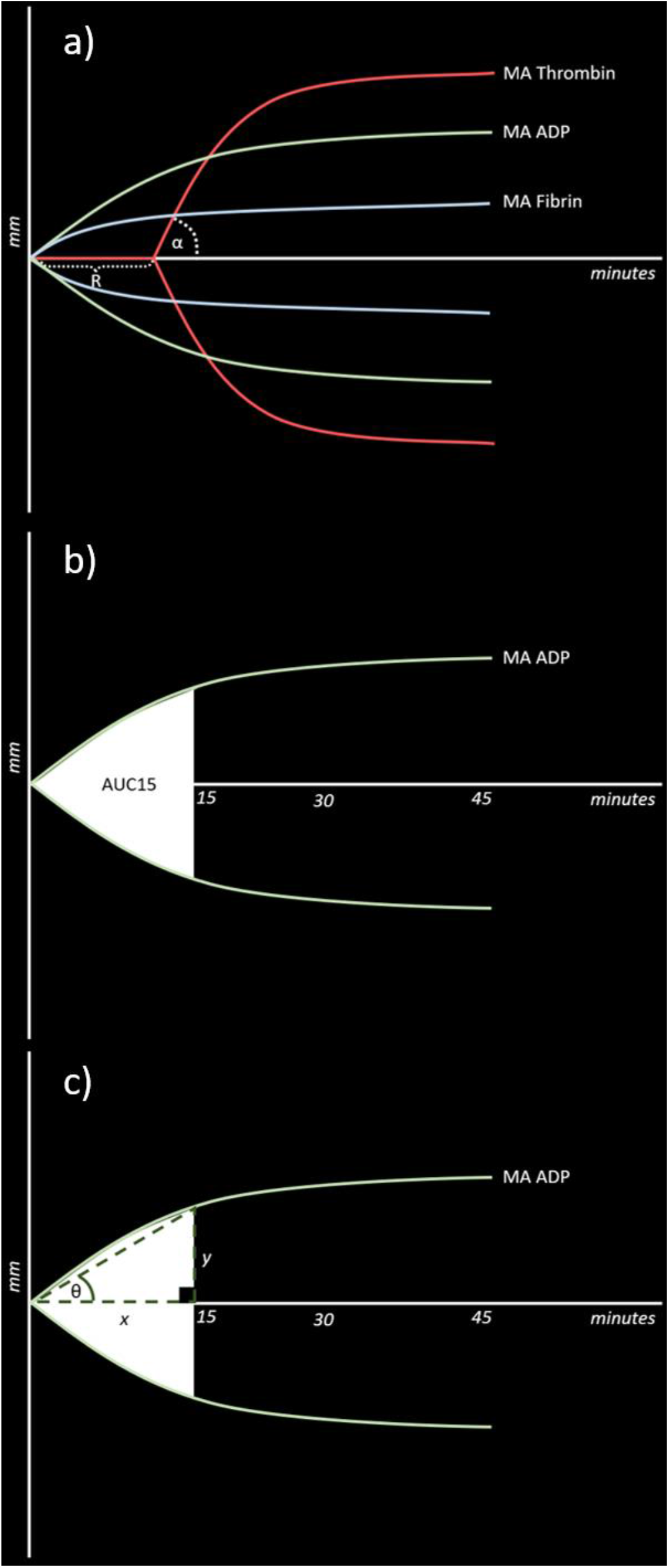
(a) An example TEG 6s tracing following a Platelet Mapping™ assay. MA = maximum amplitude, α angle = angle of thrombin curve between 2-20mm amplitude, R time = latency time between start of test until 2mm amplitude on the thrombin curve is achieved. (b) Exact area under the curve at 15 minutes that can only be calcuated using specialised computer software. (c) Right-angled triangle overlayed on AUC15 to approximate AUC15. Θ angle = an approximation of the equivalent of the α angle for the ADP curve, × = 15 minutes, y = ADP curve amplitude at 15 minutes.

The most accurate way to calculate the area under a curve is to integrate the equation for that curve between two limits as seen in Figure 1b. However, the equations for the curves produced on the TEG 6s analyser are not reported to the user. Therefore, an approximation for the area under the curve was calculated using right-angled triangles and the parameters available as seen in Figure 1c.

To calculate θ, the relative of the height of the ADP curve to the thrombin curve must be calculated as a ratio. In this example, MAs for both curves are used but in theory, any point along both curves could be used so long as the α angle has been reported. The θ angle is then calculated by multiplying the α angle by the ratio between the ADP and thrombin curves.

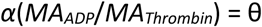

Next, using trigonometry, side length y can be represented by the equation below where y is the amplitude of the ADP curve at 15 minutes.

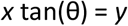

For TEG curves, the AUC is defined as both the area below the positive curve and above the negative curve. Therefore, the area of one right-angled triangle is half of the area under the curve. Using the equation for the area of a triangle, it is possible to calculate the area of one of the two right-angled triangles.

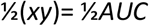

Next, substitute in a previous equation for the y-value and simplify. This is the general equation for a TEG AUC approximation at any given point in time.

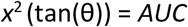

Finally, substitute in 15 minutes for × to give the following approximation for AUC15.

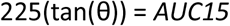

Therefore, AUC15 will be approximated for ADP curves using the equation above. Tests will be excluded where θ is ≥ 90° because a negative AUC15 would result following the use of the approximation equation. θ is typically only ≥ 90° when the amplitude of the ADP curve is substantially greater than the amplitude of the thrombin curve.

To determine if the AUC15 approximation is representative of *ex vivo* clotting response, AUC15 will be compared to the most widely used measure of *ex vivo* clotting response, the percentage of platelet aggregation.[2] The percentage of platelet aggregation is calculated using the equation below.

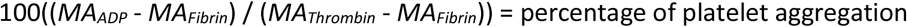

No sample size calculation was performed. Instead, all available tests stored in the internal memory of the TEG 6s analyser were retrieved for analysis to create the largest possible sample using this retrospective methodology.

Data was analysed using MedCalc^®^ for Windows Version 19.1.6.[8] AUC15 was correlated against percentage of platelet aggregation using a bivariate scatter plot with a linear regression line and 95% confidence intervals (CI) were plotted. A two-tailed Pearson correlation coefficient was also calculated. In addition, AUC15 and percentage aggregation were described using descriptive statistics.

## 3. RESULTS

In total, 47 tests were retrieved across 15 months. Two tests were excluded because θ was ≥ 90°. Therefore, 45 tests were eligible for final inclusion.

Percentage of platelet aggregation mean was 67.4% (SD: 29.1), 95% CI [58.6,76.1]. AUC15 mean was 468.4mm*min (SD: 311.4), 95% CI [374.8, 561.9].

Pearson two-tailed correlation coefficient showed that AUC15 correlated significantly with percentage of platelet aggregation (R = 0.748, 95% CI [0.582, 0.854], p < 0.001).

A bivariate scatter plot with a linear regression line and 95% CIs is plotted in Figure 2.

**Figure 2.**
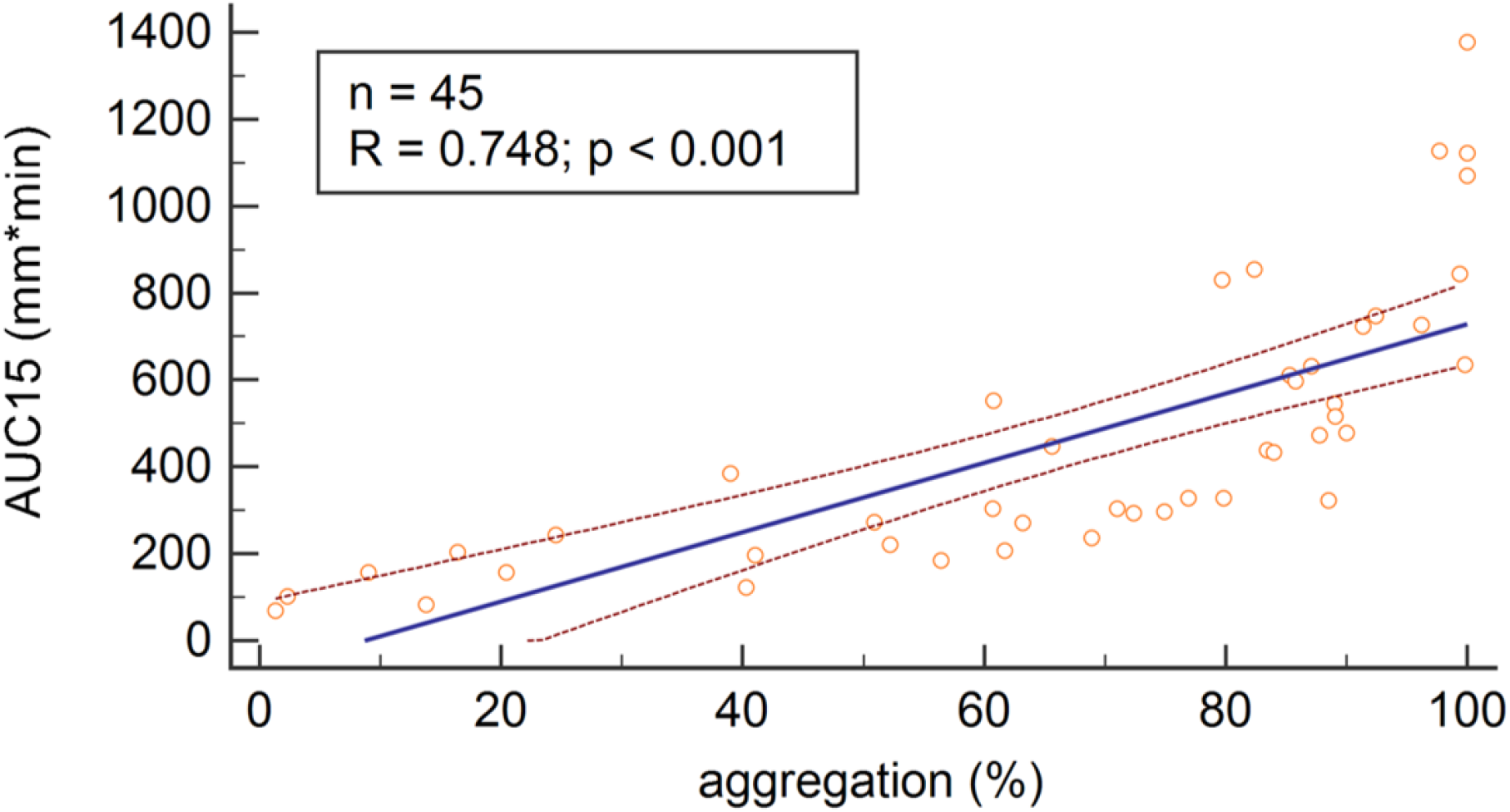
Bivariate scatter plot of percentage of platelet aggregation against AUC15 (n = 45, R = 0.748, 95% CI = [0.582, 0.854], p < 0.001). Solid blue line = linear regression line, dashed red lines = 95% CI.

## 4. DISCUSSION

This study confirms that clinicians can use parameters routinely produced by TEG analysers to approximate AUC15 for ADP curves by using the formula outlined in this study and that specialist computer software is not strictly necessary.

AUC15 remains a validated predictor of *ex vivo* clotting response to ADP receptor antagonists in healthy individuals and patients undergoing PCI. However, there have been limited studies using AUC15 as clinicians have not been able to calculate AUC15 without specialised computer software that is uploaded onto a TEG analyser. For the first time, clinicians can now approximate AUC15 using only routinely produced parameters and a calculator.

A 15-minute rapid TEG assessment of platelet function may allow surgeons to more rapidly assess a patient’s intraoperative bleeding risk who requires an emergency operation but has recently taken an ADP receptor blocker. The assessment may also allow anaesthetists to create a more informed anaesthetics management plan for such patients by making a more informed decision on whether to use a neuraxial anaesthetic technique and risk neuraxial haematoma formation.

A major limitation to this study is that all PFTs analysed were routinely performed and anonymised tests. As a result, recommended operating procedures for the TEG 6s analyser cannot be guaranteed to have been performed. However, this is unlikely to have affected the results of this study as the aim was to correlate known and calculated parameters from the same test. Therefore, the relationships between tests, and the impact of any differences that arise between tests because of different operating procedures will be minimal on the results of this study.

Another limitation is that tests which have θ ≥ 90° were excluded. As a result, not all TEG analysers will be applicable to use the formula to calculate AUC15. However, since θ ≥ 90° typically only occurred when the amplitude of the ADP curve was significantly greater than the amplitude of the thrombin curve, meaning there is very little ADP receptor inhibition on platelets, it is likely that clinicians experienced in using TEG analysers will be able to rapidly predict that the bleeding risk in these patients will not be significantly increased.

A final limitation is that not all manufactured TEG analysers report the exact same parameters. To account for these reporting differences, only parameters which are known to be widely reported on the majority of TEG analysers have been used in this study. However, it is possible that it will not be possible to use this equation for some TEG analysers.

## 5. CONCLUSIONS

This study creates the first practical formula for clinicians to approximate AUC15 of ADP curves using TEG point-of-care platelet function tests. AUC15 can allow clinicians to reliably and rapidly predict *ex vivo* platelet function at 15 minutes after the start of a test, rather than upwards of 60 minutes that traditional methods may take by using only routinely reported TEG parameters and a calculator. Our approximation of AUC15 correlates significantly with the most widely used parameter of *ex vivo* platelet function in TEG, percentage of platelet aggregation. This formula has the potential to give surgeons and anaesthetists more time to create management plans for patients who have recently taken an ADP receptor blocking antiplatelet drug who require emergency surgery or neuraxial anaesthesia. Further research employing the use of AUC15 for ADP curves in TEG should focus on the reliability of this parameter to predict clinical outcomes.

## Data Availability

Data is available upon request to the corresponding author.

## 6. GLOSSARY

*In the order that they appear in the text*

ADP: adenosine diphosphate
PFT: Platelet function tests
TEG: Thromboelastography
AUC15: Area under the curve at 15 minutes
MA: Maximum amplitude
α angle: Angle of thrombin curve between 2-20mm amplitude
R time: Latency time between start of test until 2mm amplitude on the thrombin curve is achieved
θ angle: An approximation of the α angle for the ADP curve

## ETHICAL CONSIDERATIONS

All results retrieved from the analyser were anonymised. No participants were involved in this research. Therefore, ethical approval was not required.

## ACKNOWLEDGEMENTS

There are no acknowledgements for this research.

